# A Proof of Concept for a Systems Approach to Biologically Characterize the Maternal Gut Microbiome, Immune Patterns and Mental Distress During Pregnancy

**DOI:** 10.1101/2020.11.15.20232173

**Authors:** Beatriz Penalver Bernabe, Janet L. Cunningham, Lisa Tussing-Humphreys, Ian Carroll, Samantha Meltzer-Brody, Pauline M. Maki, Jack A. Gilbert, Mary Kimmel

## Abstract

**Importance:** Screening for anxiety and depression is important but especially difficult during pregnancy. Identification and quantification of the variation of maternal biologic dimensions during the dynamic pregnancy period, e.g. immune and microbiome profiles, have the potential to greatly improve mental heath screening and patient stratification.

**Objective:** To determine if specific immune and microbiome profiles align with features of mental distress during pregnancy.

**Design:** Prospective case-control study from two cohorts in US followed from 2017 to 2019 in the second and third trimesters.

**Setting:** For the urban cohort, recruitment from an obstetric clinic; for the suburbian group, recruitment from advertisements in the hospital and surrounding communities.

**Participants:** A total of 90 pregnant women from two distinct geographic regions, with contrasting race, age, marital status, education and urban vs suburban settings.

**Exposures:** Self-reported mental health symptoms (anxiety, preceptions of stress and depression) and sociodemographic characteristics; blood was obtained for cytokine profiles and stool for microbiota analysis.

**Main Outcomes and Measures:** Dimensionality reduction clustered the symptom scores from the three self-report tools. Phenotypes based on these features were then defined.

**Results:** Factor analysis revealed four features: Burn-out, Low Self-Esteem, Mixed State, and Suicidal/Self-Harm; six phenotypes: severe, with and without suicidal thoughts, depressed with low self-steem, hyperactive, and healthy. Factors and phenotype groups were statistically associated with immune and microbiota profiles and sociodemography. The hyperactive phenotype, despite having low self-reported symptoms, had elevated inflammatory cytokines and a microbiota profile similar to the severe phenotype. Elevated T-helper cell 17 (Th17), inducer of inflammation, is related to anxiety in these cohorts. Variance in socioeconomic factors and and the gut microbiota profile can account for 50% of the variance in phenotype.

**Conclusion:** Better stratification of biological charateristics is essential for understanding complex perinatal mental disorders and can be achieved by including microbiota and immune data. Our preliminary results suggest that variance in microbial and immune factors can describe differences in mental health phenotype, and could be predictive of the women’s current or future risk for future diagnosis. These findings form testable hypotheses for larger longitudinal studies.

**KEY POINTS:** *Question:* Does self-reported mental distress in late pregnancy align with specific immune and microbiome patterns?

*Findings:* We show potential limitations in sole reliance on self-reported symptoms of mental distress in 90 subjects from two prospective cohorts. The most different groups based on symptom-based factors presented statistically similar inflammation and microbiome profiles that may reflect underreporting of mental distress symptoms.

*Meaning:* A systems biology approach identified risk immune and microbiome profiles with potential advantages to augment self-report. These biological charateristics high risk for mental distress in pregnancy should be confirmed in larger cohorts and may improve understanding of perinatal mood disorders.

## INTRODUCTION

Pregnancy is characterized by diverse biological, social, and psychological stress, stress defined as a response to demand for change.^1^ Pregnancy demands neuroendocrine, immune, social and behavioral changes, which vary across populations.^2^ While some women adapt to these changes, others’ systems over- or under-react.

Anxiety and mood shifts accompany stress responses.Two-thirds of pregnant women experience anxiety regarding fetal health, parenting, relationships, finances and career impacts.^3^ One in five meet criteria for an anxiety disorder during pregnancy.^4^ Mild to moderate levels of anxiety in the perinatal period can rapidly deteriorate, shifting towards severe mood swings and increased risk of suicide.^5^

Current screening recommendations are predominantly for depression; dominated by the Edinburgh Postnatal Depression Scale (EPDS), yet the Generalized Anxiety Disorder (GAD)-7 and Percieved Stress Scale (PSS)-14 tests have been validated during pregnancy and associate with physiological outcomes of maladaptive stress, including smaller-for-gestational-age infants.^6,7^ However, a survey of 1,500 women found that >70% minimized or completely hid their symptoms during self-reporting.^8^ Thus, unbiased, quantifiable biological measures could improve early determination of risk of maladaptive reponses and improve stratification of need of mental health support.

The microbiota-gut-brain-immune axis represents a novel biological factor associated with stress.^9–11^ The gut microbiota compositional dynamics (intestinal-associated microbes) are influenced by environment (e.g., rural versus urban, neighborhood, family) and lifestyle (e.g., diet, exercise, pet ownership).^12^ The host immune system is engaged in an intimate bi-directional interaction with the microbiota, e.g. microbial catabolism of non-digestible fibers results in short chain fatty acid synthesis mediating T-cell inflammatory suppression.^13^ Initial cross-sectional studies in pregnancy indicate an association between gut microbial composition and maternal stress and trauma.^14,15^ Animal studies in the perinatal period suggest microbial metabolism influences stress responses; and anxiety and depressive behaviors can be modified by probiotic administration.^10,16,17^ Additionally, maternal microbial composition associates with fetal neurodevelopment, and maternal stress during pregnancy influences the offsprings’ microbial composition.^10^

Here we explore perinatal mental health using systems biology approaches to integrate traditional self-report tools with microbiota and immune profiles of 90 women two sociodemographic and geographically distinct cohorts to identify biological charateristics related to mental health symptoms from. We discovered a novel set of mental distress subfactors that improve participant classification, and demonstrate statistical associations with immune patterns and microbial profiles.

## METHODS

### Participant Recruitment

Study protocols were approved by the University of Illinois at Chicago, Chicago (IRB# 2014-0325, IRB#2018-0842) and the University of North Carolina, Chapel Hill (IRB #16-0959 and 16-2783) Institutional Review Boards. Participants in Chicago were recruited during their initial obstetric visit before 16 gestational weeks (GWs). Participants in Chapel Hill were recruited through advertisement and their initial visit was before 28 GWs. Participants provided fecal samples during the second trimester (T2, range 17-28 GWs) and the third trimester (T3, range 28-40 GWs); blood samples at T2 visit. At each visit, participants completed questions about demographics and the GAD-7, PSS-14, and PHQ-9 in Chicago/EPDS in Chapel Hill.

### Factor analysis

Correlations between the Likert scores for each question of the self-reported GAD-7, PSS-14 and DEP-5 were calculated using Polychoric correlation with an oblique rotation, “Promax”. Number of factors were selected using parallel analysis. Loads for each question were estimated using unweighted least squares. Identified loads of each question were multiplied and summed up to obtain a weights for each factor per each individual. Only loads that were greater than 0.3 were employed. Finally, we normalized the weights of each factor so that they ranged between 0 to 1 and were clustered using k-means.

### Cytokine analysis

Serum samples were analyzed in duplicate using HSTCMAG28SPMX21 MILLIPLEX Human High Sensitivity T Cell 21 Plex following manufacturing instructions.

### DNA extraction and sequencing

DNA was extracted with QIAGEN® MagAttract® PowerSoil® DNA KF Kit and followed a slightly-modified manufacturing protocol,^18^ as previously described.^19^ The V4 region of the 16S rRNA gene was amplified and sequenced on a 151bp x 12bp x 151bp MiSeq run using customized sequencing primers and procedures.^19^ Empty blanks were included to control for external and cross-contamination.

### Sequence identification and filtering

DADA2^20^ was employed to establish the Exact Sequence Variants (ESV) in each sample, using default parameters unless indicated. Paired reverse and forward sequences were filtered and truncated to a maximum 150 base-pair lengths with maximum expectation of 0.75 (maxEE). After inference and chimeric removal, only sequences whose lengths were between 251 to 254 bp were used for the downstream analysis. Silva database v.132^21^ was employed to assign taxonomy to the identify ESV taxonomy. ESV coming from contamination were removed. ESV were removed that didn’t have any count above 5. Samples whose abundance was less that 1% were removed.

### Microbiome analysis

Counts were normalized using cumulative sum scaling normalization. PERMANOVA was employed to estimate differences in alpha- and beta-diversity, adjusted and stratified by possible confounders such as site, gestational age, visit, race. Associations between socio-demographics, immune, mental factors and mental phenotypes and ESVs were calculated using generalized linear models adjusted by recruitment site and research visit. Finally, for identification of the most predictive ESV and socio-demographic variables of each mental factor and each mental health phenotype, Random Forest was employed.^22^

### Statistical analysis

Unless otherwise specified, chi-square tests were used for comparisons of categorical variables, t-tests for categorial and continuous variables and Spearman correlations, and Spearman partial correlations for continuous and continuous variables. All the p-values were corrected for multiple comparisons using false discovery rate (fdr).^23^

## RESULTS

### Participant characteristics

The Chicago cohort comprised >50% Black women and ∼30% Hispanic or Latina. The Chapel Hill cohort comprised ∼80% non-Hispanic White women. Chicago and Chapel Hill participants were seen at 24 gestation weeks [range 17-28] and at 34 gestation weeks [range 28-40]. Chapel Hill participants had at least 2-years of college education, were married, and 27% obese. One third of the Chicago participants reported no college education, not married or in a committed relationship, and more than 45% obese (**Table 1**). The Chapel Hill cohort self-reported higher PSS-14 scores than Chicago participants (**Fig. 1**, p=0.003). In the 3^rd^ trimester Chapel Hill participants scored higher on the GAD-7 and DEP-5 (see methods for DEP-5 description) compared to the Chicago cohort (p=0.02).

**Table 1.** Demographic characteristics of the two different population groups. N, number; BMI, body mass index; Chicago, University of Illinois at Chicago; Chapel Hill, University of North Carolina;, false discovery rate corrected p. Gestational weeks at the V2, 24 [17,28] and at the V3 34 [28,40]. ^***^, fdr-corrected p <0.001; ^**^ fdr-corrected p <0.01; ^*^ fdr-corrected p <0.05.

|  | Chicago | Chapel Hill | Chicago | Chapel Hill |
| --- | --- | --- | --- | --- |
|  | N |  | % |  |
| Hispanic non-Black <sup>***</sup> | 13 | 1 | 28.2 | 2.1 |
| Non-Hispanic Black <sup>***</sup> | 27 | 5 | 58.7 | 10.4 |
| Non-Hispanic White <sup>**</sup> | 6 | 38 | 13.0 | 79.2 |
| Single <sup>***</sup> | 16 | 0 | 34.8 | 0 |
| Committed/Married <sup>***</sup> | 30 | 46 | 65.2 | 95.8 |
| No college <sup>***</sup> | 16 | 0 | 34.8 | 0 |
| College | 25 | 19 | 54.2 | 39.6 |
| Above college <sup>***</sup> | 5 | 15 | 10.9 | 60.5 |
| Obese<sup>*</sup> | 21 | 13 | 45.7 | 27.1 |
| Nulliparous | 13 | 20 | 29.5 | 41.7 |
|  | Average |  | Stdev |  |
| Age<sup>*</sup> | 28.4 | 32.2 | 5.5 | 2.9 |
| BMI<sup>*</sup> | 31.8 | 27.7 | 8.1 | 6.2 |

**Figure 1.**
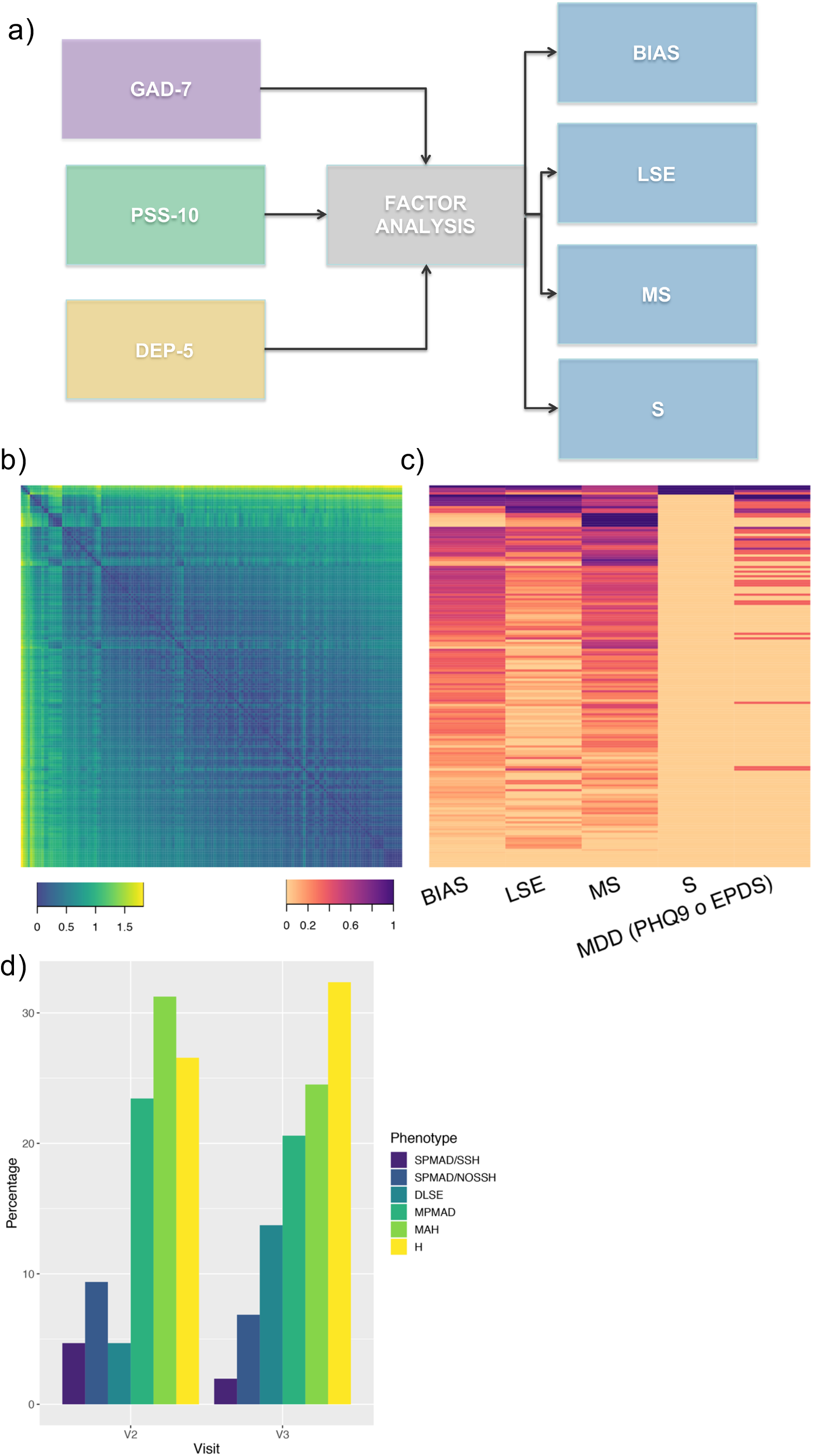
Factor analysis and participant clustering based on the scores of individual questions of GAD-7, PSS and DEP-5. a) methodology; b) clustering of total scores; c) associated values for each participant for each factor; d) percentage of participants in each cluster by trimester. No significant differences by trimester, although a trend towards significance was observed for the third phenotype “Depressed-type with lack of Confidence” (DSLE) participants (p=0.05). BIAS, burn-out, irritability, anxiety and stress; LSE, lack of confidence, low self-esteem; MS, mixed state; S, suicidal, self-harm. SPMAD/SSH: Suicidal/Self-Harm cluster; SPMAD/NOSSH: Severe PMAD without Suicidal/Self-Harm cluster; DLSE: Depressed-type with Lack of Confidence cluster; MPMAD: Moderate PMAD-more Anxious type cluster; MAH: Mild Anxiety/Healthy cluster; H: Hyperactive, less Stress/Mood Components cluster.

### Factor analysis to identify symptomic clusters into corresponding phenotypes

Factor analysis (FA) across the three screening tools (**Table S1, Fig. 2a**) resulted in four dimensions or features, some of them not previously identified, based on the most influential questions (loads) (**Table S2**): Burn-out, Low Self-Esteem, Mixed State, and Suicidal/Self-Harm. Burn-Out included questions related to coping and sense of self-control, perceived stress, worry, anxiety, and feeling on edge. FA identified Low Self-Esteem, which included questions around self-confidence to control life events. The Mixed State was most influenced by sleep disruption and a mix of anhedonia and restlessness. Suicidal/Self-Harm included only suicidal thoughts or self-harm. Using the total scores for each individual of the GAD-7, PSS and DEP-5, resulted in just two factors, Anxiety/Depression and Perceived Stress (**Fig. S1**). Because DEP-5 included questions that differed slightly depending on EPDS or PHQ-9, the FA was also completed without the DEP-5, and showed no influence on the factors. There were some differences seen when the FA was done for each site alone, but still resulted in three groups for each site that included predominant components of Burn-out, Mixed state, and Low Self-Esteem (**Table S3**).

**Figure 2.**
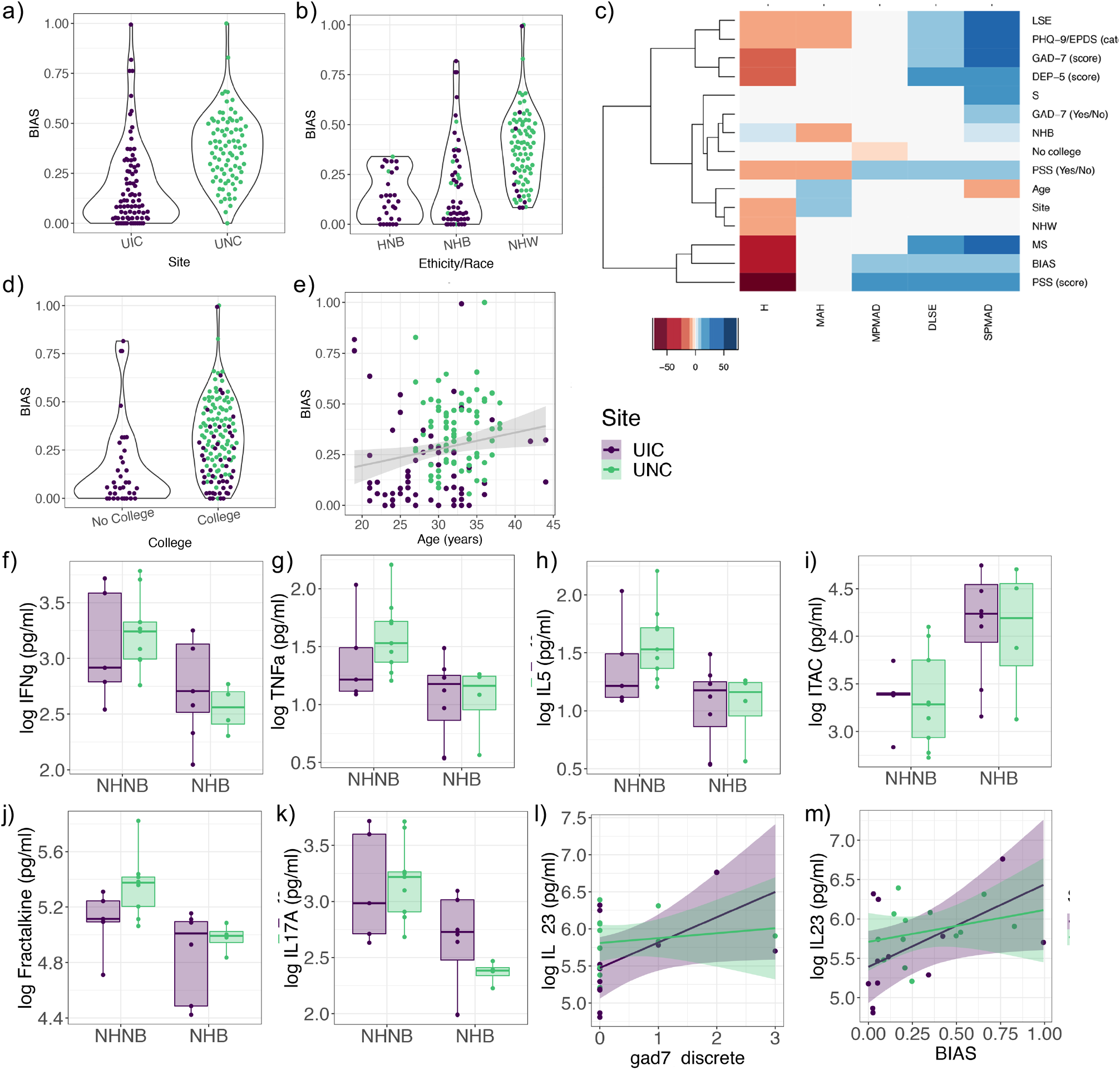
Associations between participant demographic characteristics, mental factors and phenotypes and cytokine profiles. Out of all possible associations, BIAS factor was associated with some of the sample characteristics (p<0.001), including a) recruitment site, b) ethnicity and race, c) college achievement and d) age. Cluster mental health phenotypes were also associated with several self-reported questionnaires and socio-demographics (e). Out of all the 21 panel of cytokines that were measured in serum in the second trimester, only those that were significantly associated with ethnicity, race, education, age, parity or site, are presented here (p<0.05). Similarly, only those cytokine concentrations in serum that were associated with stress, anxiety, depression or their corresponding factors, are shown here. HNB, Hispanic non-Black; NHB, non-Hispanic Black; NHW, non-Hispanic White. SPMAD: Severe PMAD with and without Suicidal/Self-Harm; DLSE: Depressed-type with Lack of Confidence; MPMAD: Moderate PMAD-more Anxious type; MAH: Mild Anxiety/Healthy; H: Hyperactive, less Stress/Mood Components.

Based on individual variation in the features, six phenotypic clusters were identified (**Fig. 1b-c**): (i) severe depression and anxiety with suicidal ideation and/or self-harm; (ii) severe depression and anxiety without suicidal ideation and/or self-harm; (iii) Moderate Anxiety; (iv) low self-esteem and depression (Depressed); (v) mild anxiety (Healthy); and (vi) hyperactive with less anxiety and depression components (Hyperactive). Groups 1 and 2 had the fewest participants and since suicidal ideation and/or self-harm was only disclosed in the Chapel Hill cohort with low symptom severity (1 in the scale of 0-3), both severe groups were combined: Severe. No significant differences between the groups in the second and third trimesters were observed (p>0.01) (**Fig. 2d**).

Burn-Out was associated with site (Chapel Hill), age, ethnicity and race, and college education (**Fig. 2a-d**; frd-corrected p<0.001). Women in the Severe cluster (**Fig. 2e**) were younger and more likely self-reported as non-Hispanic Black (NHB). In the Moderate Anxiety cluster, participants had higher levels of Burn-Out and tended to have higher education. Participants belonging to the Hyperactive cluster self-reported as NHB and self-reported lower levels in all questionnaires and mental factors. Women in the Healthy group, were older, more confident in their capabilities, and mostly from the Chapel Hill cohort.

### Cytokines patterns

NHB women had greater concentrations of serum ITAC (**Fig. 2a**) and lower Fractalkine, IL-17, IFN-gamma, TNF-alpha, and IL5 than NHB women (**Fig. 2b-f**). GAD-7 and Burn-Out factor scores were positively correlated with IL-23, and more pronounced in the Chicago cohort (**Fig. 2g, h**). Most cytokines were positively correlated with each other except ITAC was negatively associated with MIP-3a and Fractalkine. IL-10, IL-5 and IL-7 did not clearly belong to a specific cluster, yet IL-10 was positively associated with IFN-g or IL-12 (p70) and IL-5 was associated with TNF-a (**Fig. 3a**, rho>0.9, fdr-corrected p<0.001). Participants from the same phenotypic group or ethnicity/race didn’t cluster together (**Fig. 3b**) and the Hyperactive group was split into those with the highest and lowest levels of cytokines (**Fig. 3c)**. Compared with the Healthy cluster, when looking at the entire Severe and Hyperactive groups, they presented with opposite directions in inflammatory levels, while the Moderate Anxiety and Depressed groups were similar to Healthy cluster (**Fig. 3d**, fdr-corrected p<0.05 for all the above comparisons).

**Figure 3.**
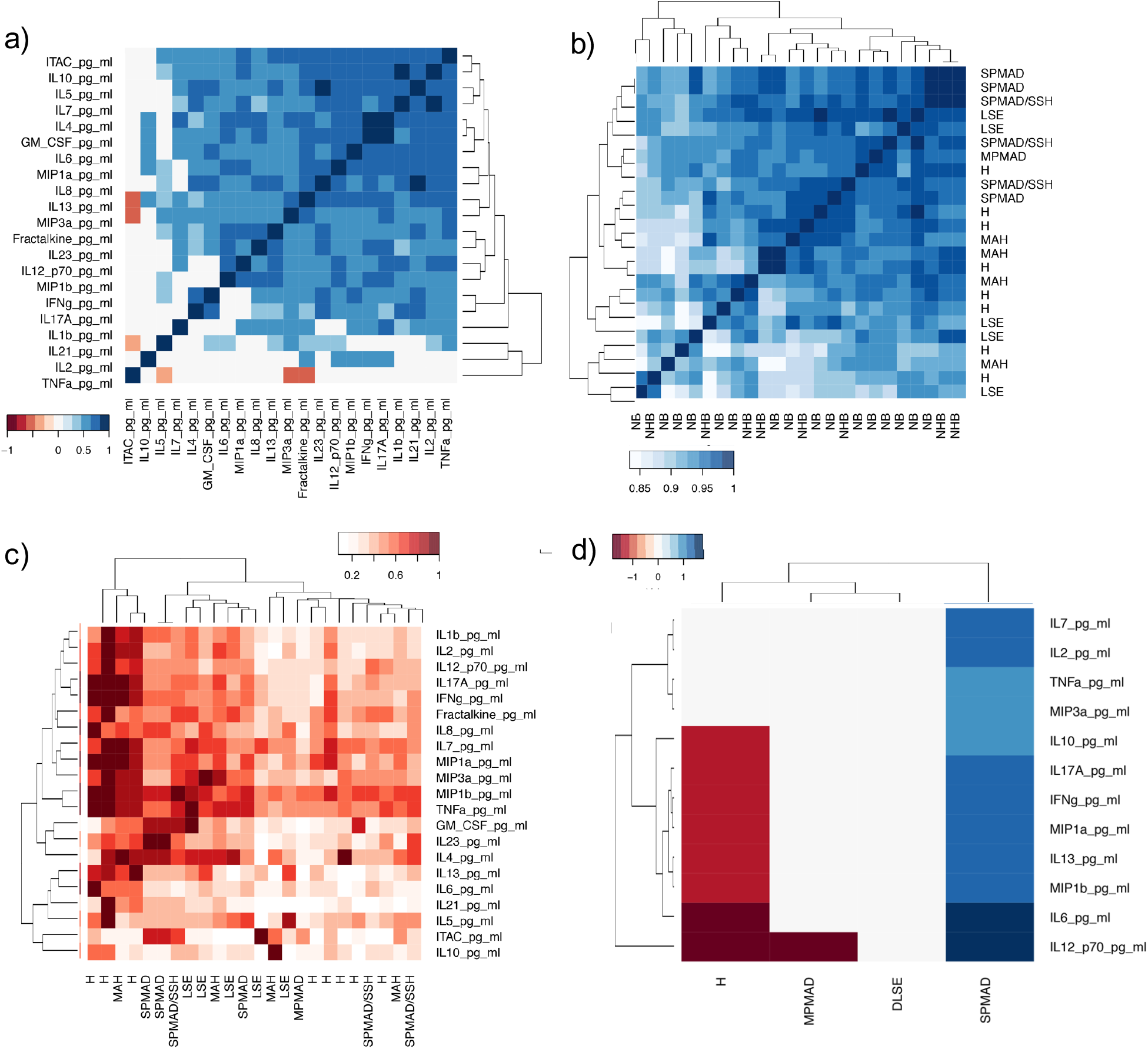
Significant associations between cytokines and phenotypes. a) correlations between cytokines abundance (fdr-corrected p<0.001); b) Correlation between participants cytokines; c) normalized cytokine abundance as a function of participant phenotype; d) Association between mental health phenotypes with respect to those with mild anxiety/healthy and normalized abundance of significant cytokines (fdr-corrected p<0.05). SPMAD/SSH: Suicidal/Self-Harm; SPMAD: Severe PMAD without Suicidal/Self-Harm; LSE: Depressed-type with Lack of Confidence; MPMAD: Moderate PMAD-more Anxious type; MAH: Mild Anxiety/Healthy; H: Hyperactive, less Stress/Mood Components; NB: non-black; NHB: non-Hispanic black.

### Microbiome patterns and mental health factors

Alpha- and beta-diversity estimates had no significant correlation with any of the socio-demographic variables, gestational timepoints, body mass index, self-reported GAD-7, PSS-14 or PHQ-9/EPDS (binary) values, and only limited correlation with some of the immune markers (**Fig.S2 and S3)**. Several ESVs were significantly associated with the Burn-Out, Mixed State and Low Self-Esteem factors (fdr-corrected p<0.001, **Fig. 4a**). Only two ESVs connected the 3 factors, Burn-Out, Mixed State and Low Self-Esteem factors: *Lactobacilus inners* and the genus *Prevotella*. Increased proportions of these two ESVs was associated with higher levels of Burn-Out, Low Self-Esteem, and Mixed State. The Mixed State negatively associated with the proportion of *Bifidobacterium bifidum, Senegalimassilia anerobia, f*amilies Clostridiales Incertae Sedis Family XIII, Lachnospiraceae, two members of the Ruminococaceae family, and the genera *Porphyromonas* and *Sneathia* ESVs (**Fig. 4a**). Burn-Out was positively correlated with the proportion of *Streptococcus mutans*, and negatively correlated with *Peptoniphilus* and *Parabacteroides goldsteinii*. The predicitive capabilities of the socio-demographic and relative abundance of the significant ESVs to explain the variabilities in the mental health factors were low (Burn-Out 17%, Low Self-Esteem 19% and Mixed State 1%) (**Table S4**). Interactions between cytokine concentrations in serum and ESV relative abundance was mostly inversely correlated, with IL-23 as one of the keystone cytokines that interacted with the greatest numbers of ESVs, followed by IL-10 (**Fig. 4b**).

**Figure 4.**
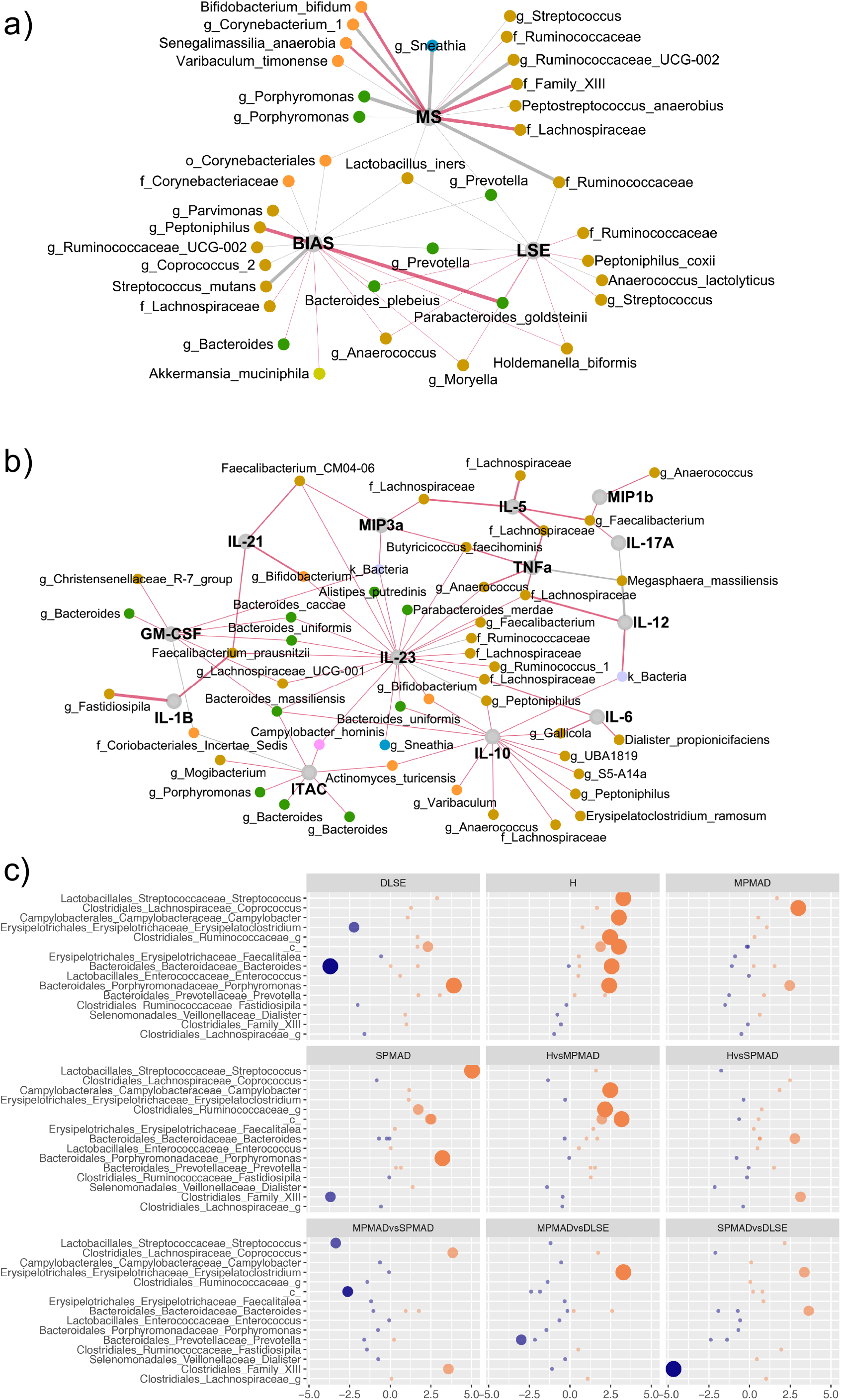
Significant associations (fdr-corrected p<0.001) between ESVs, mental factors cytokines and mental health clusters. a) ESVs and mental factors (fdr-corrected p<0.001) between. b) ESVs and cytokines (fdr-corrected p<0.01). Node colors are associated with their phylum; red edges between nodes represent a negative interaction; grey edges represent a positive interaction. Edge thickness is associated with the weight/strength of the association. C) Fold change differences in the normalized abundance of the significant ESV between the all five identified mental health phenotypes. Size is inversely associated with the fdr-corrected p-value for each of the ESV for a given comparison. Orange circles indicate positive fold-changes; blue circles indicate negative fold-change. Only ESV that were significance at a given comparison are depleted here. SPMAD, severe depression and anxiety with/without suicidal ideation and/or self-harm; MPMAD, Moderate Anxiety; DLSE low self-esteem and depression (Depressed); H, hyperactive with less anxiety and depression components (Hyperactive)

### Microbiome patterns and phenotypes

Alpha- and beta-diversity was not significantly different between phenotype clusters, but 15 ESVs had differential proportions (**Fig. 4c)**. The Hyperactive and Healthy clusters were the most dissimilar, while Hyperactive and Severe clusters were the most similar. Random Forest models indicated that sociodemographic and ESV proportions could partially predict the mental health cluster against the healthy control (**Table S5**). Random forest could predict the hyperactive phenotype best (Area under the curve, AUC 72%), followed by the combined Severe and Moderate Anxiety (AUC 63%) and Depressed (AUC 59%) clusters. Age at recruitment and the recruitment site were among the top predictors for all the groups and the most predictive variables for the Depressed phenothype. *Faecalibacterium* and Lachnospiraceae ESV proportions predicted the Severe/Moderate Depression and Anxiety. While the validated Random Forest model could predict with good accuracy the participants with Severe and Moderate Anxiety, the model had an inferior performance predicting controls, with more than 60% of controls predicted as participants with the Severe and Moderate Anxiety combined group.

### DISCUSSION

Combining cohorts of women with two distinct sociodemographic characteristics and geographies underdscored the limitation of current diagnoses in diverse populations. Using a combination of statistical and computational methods, a novel set of mentral distress subfactors enabling more detailed phenotyping than traditional approaches (e.g., healthy, anxious, stressed, anxious and stressed) were identified to compare with immunologic and microbiome profiles. This also highlighted the limitation of relying on self-reported data alone. Our results point to novel data-driven testable hypotheses to explore in on-going studies.

Low-self esteem is a risk factor for perinatal depression or anxiety,^24^ but our study is the first to describe how symptoms of low-self esteem predominate as a form of moderate depression. Burn-out is not only prevalent in mothers with children of all ages, but also during pregnancy, as demonstrated. Sleep disturbances in pregnancy are associated with the development of manic and depressed symptoms postpartum; therefore it is important that we identified an antenatal factor of restlessness and anhedonia related to sleep disruption, as it may be a precursor to manic and depressed symptoms postpartum.^25,26^

DSM diagnoses are often limited and can contribute to misdiagnosis, particularly in younger ethnic minorities.^27^ In addition, the data-driven Woods-Giscombe’s Superwoman Schema might explain our observations related to race. Woods-Giscombe proposed that Black women are more likely to feel an obligation to manifest strength and independence, suppress emotions, succeed despite limited resources, and help others.^28^ Yet, these expectations and chronic stress such dealing with racism associate with emotional eating, dysfunctional sleep patterns, and postponement of self-care.^28^ Social factors such as low education, negative life events, ethnic-minority status, and ambivalence about the pregnancy are predictors of high depressive symptom burden. ^29^ Even though the Chicago cohort had a greater number risk factors, the number participants reporting low or no anxiety or depression was significantly lower than the Chapel Hill cohort. The Hyperactive group was composed primarily of younger and Black women. It is plausible that some of the women clustered in this group partially or not at all disclosed their mental distress; explaining how the Hyperactive was most similar to the ‘Severe’ group in microbial composition patterns and had some indiviudals with higher cytokine concentrations.

The results presented here allow us to posit several key hypotheses:

### Hypothesis 1: Increased prevalence of T-helper 17 immune cells and gut microbal dysbiosis correlate with pregnancy-associated anxiety

IL-23, a central hub in the immune-microbia network (**Fig. 4b**), was signifcantly positively associated with GAD-7 and the Burn-Out factor (**Fig. 2l,m**). IL-23 directs the development and maturation of T-helper 17 (Th17) cells, producers of IL-17. IL-23 also stimulates innate immune cells to increase IL-6 production that leads to less T-regulatory immune cells (Treg).^30^ Osborne, Brar, and Klein propose that dysregulation of the ratio between Th17 and Treg in pregnancy may contribute to anxiety and depression.^30^ Th17 regulated pathways are prevalent in populations with severe chronic fatigue and Th17 related pathways are considered to be the mediators to the chronic disease.^31,32^ Thus, Th17 may serve as transdiagnostic marker of poor stress adaptation and for intervention. ^32^ Th17 cells control and are controlled in part by commensal microorganisms.^33^ Increased levels of cytokines IL-21 and IL-23 and Mixed State factor were associated with lower relative counts of *Bifidobacterium bifidum*. Whole grain barley diets increase the proportion of *Bifidobacterium* ^34^, which suggests diet as intervention. Importantly, this maternal shift may also transmit a long-lasting shift in Th17 to the infant, increasing the risk for schizophrenia.^35^

### Hypothesis 2: Integration of immune, microbial and sociodemographic information will identify stress responses associated with negative pregnancy and mental health outcomes for mother and child; regardless of self-report of mental distress

The immune system is extraordinarily dynamic over pregnancy and its trajectory is modifiable by socio-demographic factors.^36^ For example, Corwin, et. al., hypothesized, a glucocorticoid resistance model in low-income minority women could explain higher cortisol levels, an exaggeration of normal immunomodulation, in the third trimester, without any significant changes in pro-inflammatory cytokines.^37^ Associations between cytokines and perinatal depression and anxiety are inconsistent. Some authors report negative associations between depressive symptoms in pregnancy and inflammatory cytokines (e.g., IL-1b, TNF-a, IL-7 studied in the second trimester; TRAIL, M-CSF, Fractalkine studied at 38 weeks gestation or at delivery); while others have reported positive associations between depressive and anxiety symptoms in the third trimester with cytokines IL-6, IL-15, and CCL3.^36^ While there might be multiple reasons, host-microbial interactions provide important additional information. For instance, *Faecalibacterium* has been previously associated with lower levels of inflammation,^38^ and was an important predictor of the Severe phenotype. *Megasphaera*, positively associated with multiple cytokines including IL-17A, IL-12, TNF-alpha in our study, has been associated with increasing risk of preterm birth, ^39^ possible shared underpinnings of antenatal depression and preterm birth.^40^ Given that socio-demographic factors are predictors of the Hyperactive phenotype, modeling must integrate all these dimensions, i.e., socio-demographic, immune and microbial. Age may represent complex and confounding interactions, such as stigma or difficulties identifying and communicating mental distress.

## Strengths and Limitations

While our study has several limitations including small sample size, recruitment methods, lack of past psychiatric history, no diagnosis by mental health professionals, only two perinatal visits, our study is the largest to date to integrate socio-demophics and host-microbial interactions during pregnancy. Hower, understanding the dynamic host-microbial interactions requires larger, densly sampled, longitudinal studies in which mother-dyads could serve as their own controls. Nevertheless, our study outlines a novel use of statistical and computational tools to ascertain not only new mental health dimensions, but also immune and microbiome patterns.

## CONCLUSIONS

Our study provides the largest sample size and most diverse participants to date assessing mental distress in relation to the immune response and microbial composition. Our preliminary findings suggest that traditional psychiatric phenotypes do not necessarily align with biological measures, underscoring the essentiality to include microbial and immune patterns in larger cohorts of mother-child dyads alongside newly defined factors. These novel approaches could be more indicative of the experiences of women in the perinatal period and open the door closer towards precision medicine and novel interventions.

## Data Availability

Raw data have been deposited in SRA BioProject PRJNA667109

## Data availability

Raw data have been deposited in SRA BioProject PRJNA667109

## Acknowledge

BPB was funded by the Arnold O. Beckman Postdoctoral Award, K-12 Biomedical Interdisciplinary Research in Women’s Helth Award. MK is supported by the NIMH K23 Training Award (1 K23 MH110660-01) and her research was also funded through a NARSAD Young Investigator Award from the Brain and Behavior Research Foundation and the P&S Fund. This work has been funded by the NICHD 5R03HD095056-02. REDCap application is supported though the Center for Clinical and Translational Science (CCTS) UL1TR002003.

