## Supplementary Information for "A Proof of Concept for a Systems Approach to Biologically Characterize the Maternal Gut Microbiome, Immune Patterns and Mental Distress During Pregnancy"

**SUPPLEMENTAL METHODS**

**Participant Recruitment**

Study protocols were approved by the University of Illinois at Chicago (IRB# 2014-0325, IRB#2018-0842) and the University of North Carolina (IRB #) Institutional Review Boards. We recruit a total of 46 from the University of Illinois at Chicago and 44 from the University of North Carolina. Participants from the University of Illinois at Chicago (Chicago) were recruited before 16 gestational weeks during their initial obstetric visit for the current pregnancy. Participants from the University of North Carolina (Chapel Hill) were recruited through advertisement and their initial visit was anywhere before 24 gestational weeks. Participants provided stool samples or rectal swabs at the visit during their second trimester (T2, range 17-28 gestational weeks) and at the visit during their third trimester (T3, range 28-40 gestational weeks) and blood samples in their T2 visit. Rectal swabs at Chicago were collected during the participant visit. Most of the stool samples from Chicago and all the stool samples from the Chapel Hill were collected at home. Chicago participants preferentially shipped the samples to Chicago; some Chicago and all Chapel Hill participants brought their specimens at their clinical or study visit. Samples shipped to Chicago were previously frozen below 32 F before depositing them for shipping. Thus, some samples remained at room temperature for more than 24 hours prior arrival to Chicago. Stool samples from the Chapel Hill cohort were collected by participants at home, put on ice and transported within 24 to 48 hours. Fecal specimens from both locations were homogenized, aliquoted, and froze at -80 C by research coordinators. Participants complete a series of questionnaires about the levels of anxiety (using GAD-7), stress (PSS) and depression symptoms using PHQ-9 in Chicago and EPDS at Chapel Hill, and demographics. Researchers had access to their electronic medical records. Gestational weeks and complications during pregnancy were obtained from them.

**Factor analysis**

Factor identification was performed in the following manner. First, we calculate the correlation between the Likert scores for each questions of the GAD-7, PSS-14 and DEP-5 self-reported questionnaires. As anxiety, stress and depression are highly correlated, we employed polychoric correlation coefficients between the scores with an oblique rotation, “Promax”. Number of factors were selected using parallel analysis that compares the scree of factors of the observed data with that of a random data matrix of the same size as the original. The value was selected for the number of factors in which there was the minimum difference between the eigen values of the raw data and of the resample data. Loads for each question from the 3 self-reported questionnaires was estimated using unweighted least squares solution with 2,000 iterations and correlation preserving approach to calculate the factor scores ("*tenBerge*"). Then we multiplied the loads of each questions for the Likert scores for each interview and we summed up the results to obtain a weights for each factor per each individual. Only loads that were greater than 0.3 were employed. Finally, we normalized the weights of each factor so that they ranged between 0 to 1. Using the clustering algorithm k-means of the scaled weights between 0 and 1, we established the number of groups that the participants could be split into. All the computations were performed in R using the “*psych*” package.^1^ Only participants that had complete datasets, i.e., PSS, PHQ-9 or EPDS, and GAD-7 were included in the factor analysis.

**Cytokine analysis**

Serum samples were analyzed in duplicate using HSTCMAG28SPMX21 MILLIPLEX Human High Sensitivity T Cell 21 Plex following manufacturing instructions. HSTCMAG28SPMX21 can simultaneously analyzes Fractalkine, GM-CSF, IFNγ, IL-1β, IL-2, IL-4, IL-5, IL-6, IL-7, IL-8, IL-10, IL-12 (p70), IL-13, IL-17A, IL-21, IL-23, ITAC, MIP-1α, MIP-1β, MIP-3α, TNF-α

**DNA extraction and sequencing**

DNA was extracted with QIAGEN® MagAttract® PowerSoil® DNA KF Kit and followed a slightly-modified manufacturing protocol.^2^ As, previously described,^3^ we amplified the V4 region of the 16S rRNA gene of the barcoded fecal samples with region-specific primers, 515F-806R, that include adapter sequences (amplicons). After PCR amplification, we employed PicoGreen (Invitrogen) to quantify the amplicons that were pooled together to reach an equimolar mixture. Then, the pool sample was sequenced on a 151bp x 12bp x 151bp MiSeq run using customized sequencing primers and procedures.^3^ Three type of empty blanks were also included in the sequencing: i) clean rectal swabs to identify possible contamination in the swabs, n=16; ii) open rectal swabs exposed to the clean hood environment during the extraction and barcoding protocol, n=16; iii) empty vials to account for the possible contamination present in the extraction kits and downstream processing, n=35.

**Sequence identification and filtering.**

DADA2^4^ was employed to established the Exact Sequence Variants (ESV) present in each sample, using default parameters unless indicated otherwise. Illumina forward and reverse sequences were split by sample using QIIME.^3^ Paired reverse and forward sequences were filtered and both truncated to a maximum 150 base-pair lengths with maximum expectation of 0.75 (maxEE). Out of the 221 samples, eleven samples were removed due to low number of reads (<5 reads). One sample was a empty vial sample and 6 of them belonged to the North Carolina cohort. After inference and chimeric removal, only sequences whose lengths were between 251 to 254 bp were used for the downstream analysis. Silva database v.132 (12) was employed to assign taxonomy to the identify ESV taxonomy. We observed sample cross-contamination (**Fig. S7**), some fecal matter contaminated the controls blanks (i.e., clean rectal swabs, environmental-exposed rectal swabs, and empty vials). To account for cross-contamination and identify only the ESV present in the control blanks, we assumed that ESVs that were not present in at least 90% of the samples should be removed (**Fig. S8**) based on the ESV prevalence in the sample. Additionally, we removed ESV that didn’t have any count above 5. Finally, samples whose abundance was less that 1% were further removed. All analyses were done in R.^5^

**Microbiome analysis**

Counts were normalized using cumulative sum scaling normalization (CSS) and relative abundance was calculated using the CSS normalized data.^6^ Alpha- and beta-diversity are calculated with the package *phyloseq*^7^ in R. For Unifrac^8^ calculations, data were not CSS-normalized and rarefaction was set to 5,000. For determination of statistically significant differences in alpha-diversity, we employed PERMANOVA and adjusted and stratified by possible confounders such as site, gestational age, visit, race. We included socio-demographics, immune and mental factors and mental phenotypes to determine their associations with sample alpha-diversity. Similarly, for beta-diversity associations with socio-demographics, immune and mental factors and mental phenotypes PERMANOVA tests adjusted by the recruitment side were used. Associations between socio-demographics, immune and mental factors and mental phenotypes were and ESVs were calculated using generalized linear models adjusted by recruitment site and research visit using the R package *metagenomeSeq*.^9^ Finally, for identification of the most predictive ESV and socio-demographic variables of each mental factor and each mental health phenotype, we employed Random Forest.^10^ We employed 70% of the data to train the model and 30% as a test set. Random Forest parameters were initially selected based on the combination of number of trees, number of variables randomly sampled as candidates at each split, size of sample to draw and minimum size of terminal nodes that produce the minimum model error. To avoid overfitting, we removed all the variables that didn’t reduce the mode error in order of importance. The random forest model was again trained with this smaller variable subset.

**Statistical analysis**

Unless specified otherwise, comparison between categorical variables were performed with chi-square tests, and between categorial and continuous and continuous-continuous variables, we employed t-tests, Spearman correlations and Spearman partial correlations. Unless stated otherwise, all the p-values were corrected for multiple comparisons using false discovery rate (fdr).^11^ Analysis was conducted in R^5^ and figures were produced using the package *ggplot2*.^12^

**SUPPLEMENTARY INFORMATION**

**Table S1. Depression questionnaire (DEP-5) based on common questions between PHQ-9 and EPDS**

| **DEP-5** | **PHQ-9** | **EPDS-10** |
| --- | --- | --- |
| 1 | Little interest or pleasure in doing things | I have looked forward with enjoyment to things |
| 2 | Feeling bad about yourself or that you are a failure or have let yourself or your family down | I have blamed myself unnecessarily when things went wrong |
| 3 | Trouble falling or staying asleep, or sleeping too much | I have been so unhappy that I have had difficulty sleeping |
| 4 | Feeling down, depressed, or hopeless | I have felt sad or miserable |
| 5 | Thoughts that you would be better off dead, or of hurting yourself | The thought of harming myself has occurred to me |

**Table S2. Loading for each of the identified factors using total scores for GAD-7, PSS and DEP-5; of using individual questions from all the three questionnaires.** Between parenthesis it is the questionnaire and question number where each item belongs to**.**

| Factor | Questionnaire/Specific question | Load |
| --- | --- | --- |
| BURN-OUT/IRRITABILITY, ANXIETY, STRESS | In the last month, how often have you found you could not cope with all the thing that you had to do? (PSS-6) | 0.88 |
|  | In the last month, how often have you been angered because of things that were outside of your control? (PSS-9) | 0.85 |
|  | In the last month, how often have your felt that you were unable to control important things in your life? (PSS-2) | 0.82 |
|  | In the last month, how often have you felt nervous or “stressed”? (PSS-3) | 0.78 |
|  | Feeling bad about yourself or that you are a failure or have let yourself or your family down/ I have blamed myself unnecessarily when things went wrong (DEP5-4) | 0.76 |
|  | In the last month, how often have you felt difficulties were piling up so high that you could not overcome them? (PSS-10) | 0.74 |
|  | In the last month, how often have you been upset because of something that happened unexpectedly?  (PSS-1) | 0.72 |
|  | Feeling afraid as if something awful might happen (GAD7-7) | 0.60 |
|  | Becoming easily annoyed or irritable (GAD7-6) | 0.55 |
|  | Feeling nervous, anxious, or on edge (GAD7-1) | 0.50 |
|  | Feeling down, depressed, or hopeless/ I have felt sad or miserable (DEP5-2) | 0.47 |
| LACK OF CONFIDENCE/LOW SELF-ESTEEM | In the last month, how often have you felt that you were on top of things? (PSS-4) | 0.89 |
|  | In the last month, how often have you felt confident about your ability to handle your personal problems? (PSS-8) | 0.89 |
|  | In the last month, how often have you been able to control irritations in your life? (PSS-5) | 0.79 |
|  | In the last month, how often have you felt that things were going your way? (PSS-7) | 0.77 |
| MIXED STATE | Trouble falling or staying asleep, or sleeping too much/ I have been so unhappy that I have had difficulty sleeping (DEP5-3) | 0.90 |
|  | Little interest or pleasure in doing things/ I have looked forward with enjoyment to things (DEP5-1) | 0.63 |
|  | Being so restless that it's hard to sit still (GAD7-5) | 0.62 |
|  | Worrying too much about different things (GAD7-3) | 0.57 |
|  | Trouble relaxing (GAD7-4) | 0.57 |
|  | Not being able to stop or control worrying (GAD7-2) | 0.52 |
| SUICIDAL/SELF-HARM | Thoughts that you would be better off dead, or of hurting yourself/ The thought of harming myself has occurred to me (DEP5-5) | 0.81 |

**Table S3. Factor analysis variations as a function of self-reported questionnaires employed and stratification by site.** Results using all the available data and questionnaires (“*All Data*”); all available data and just GAD-7 and PSS questionnaires (“*All data without DEP-5*”); all questionnaires with just Chicago data (“*Data Only Chicago*”); all questionnaires with just North Carolina data (“*Data Only Chapel Hill*”). Cell color are associated with the load similarity to the factor analysis results obtained with all the available data (“*All Data*”). Green colors indicated an agreement between the two factor analysis; orange indicate that the results do not agree for the specific question and the loads were greater than 0.6; yellow indicate that the results do not agree for the specific question and the loads were less than 0.6.

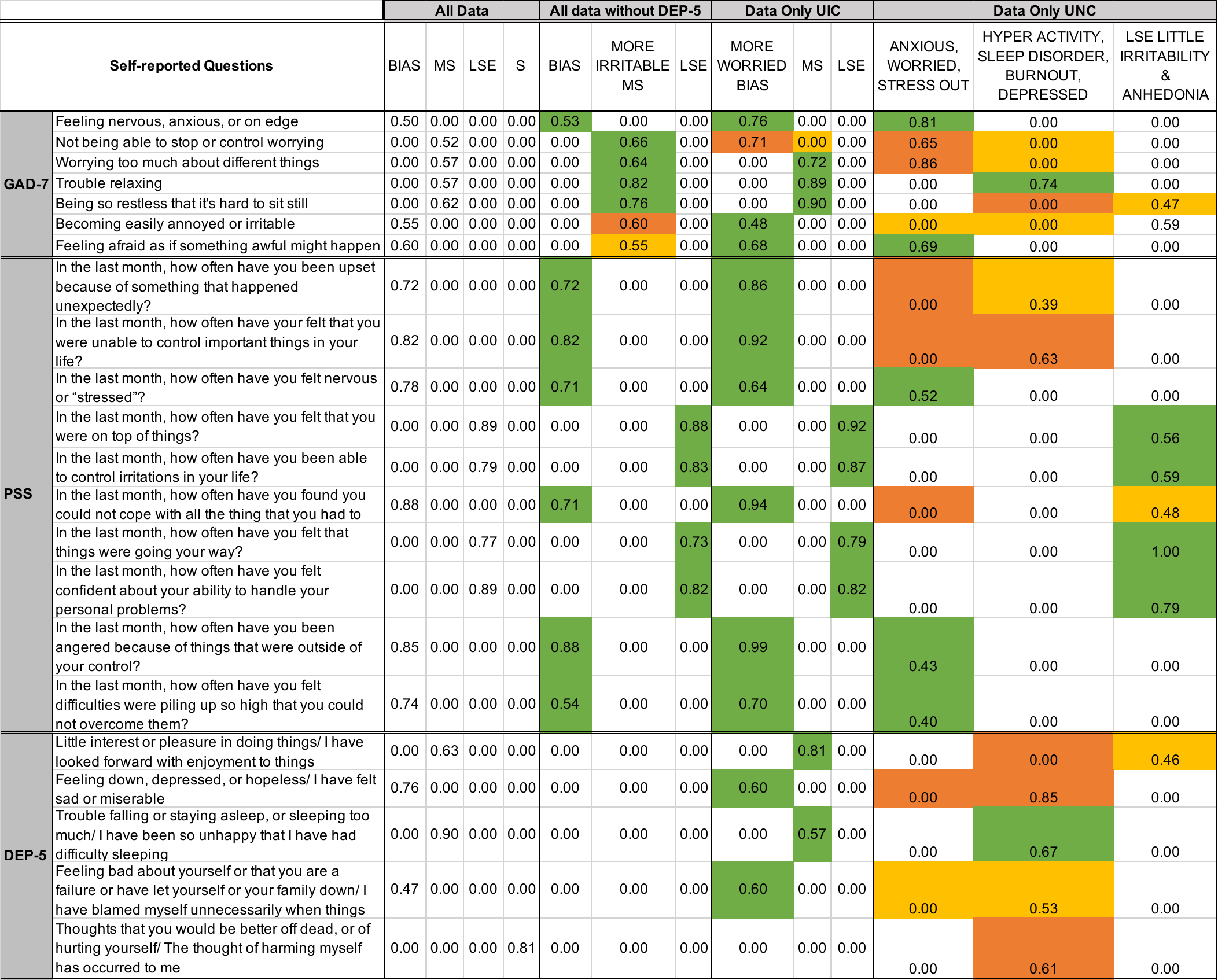

**Table S4. Socio-demographics and ESV relative abundance predictors of BIAS, LSE and MS mental health factors.** Predictions were obtained with an optimized Random Forest model (see methods for more details)

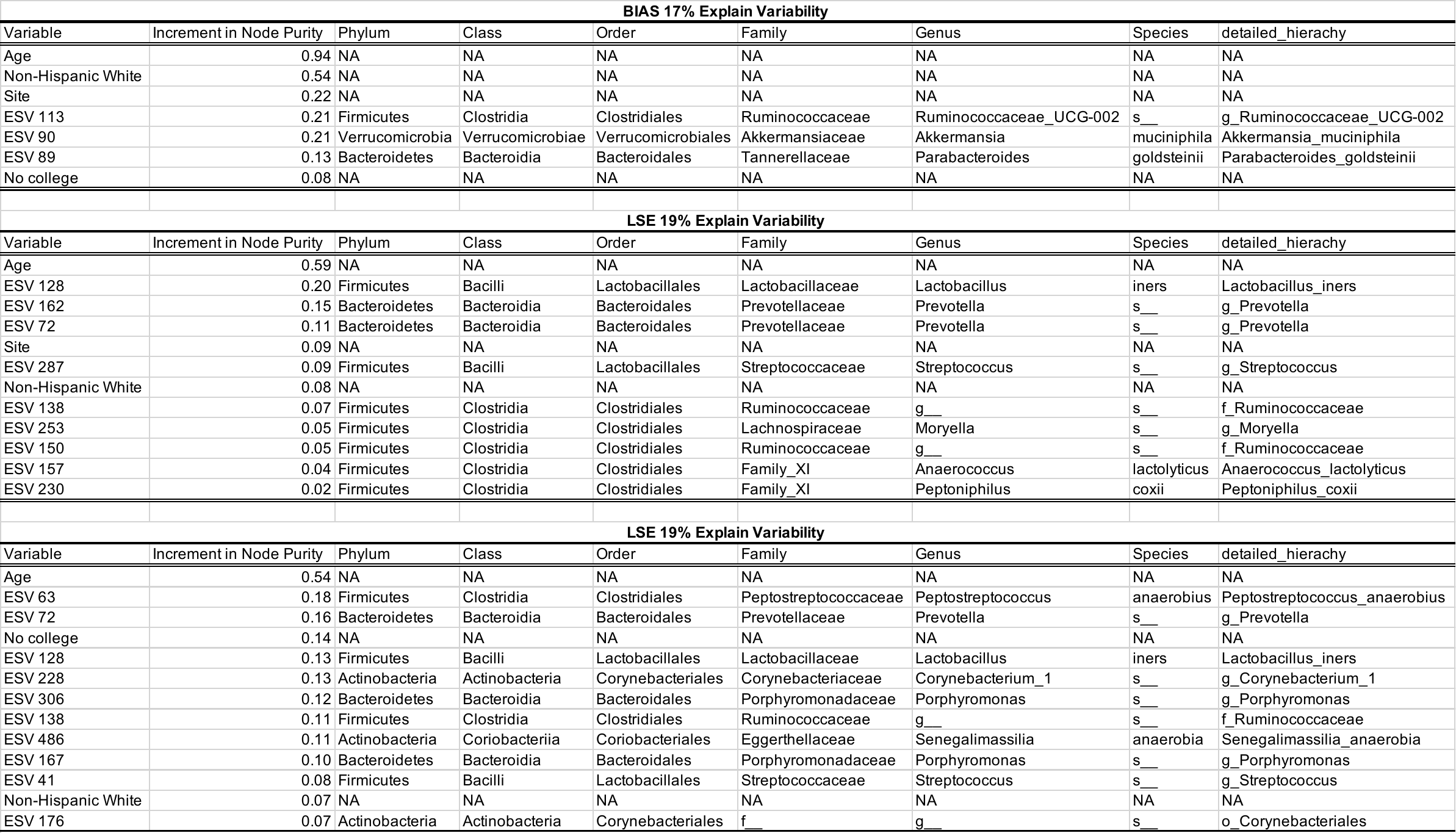

**Table S5. Socio-demographics and ESV relative abundance predictors of the mental healh phenotypes.** We have combine the Severe and Moderate cases to include the number of cases. Predictions were obtained with an optimized Random Forest model (see methods for more details)

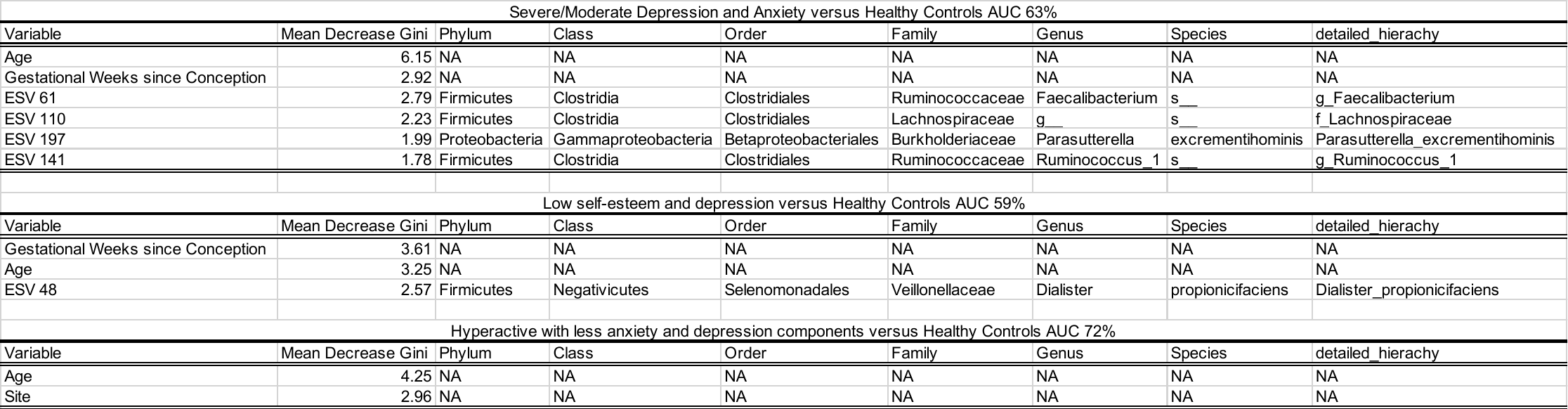

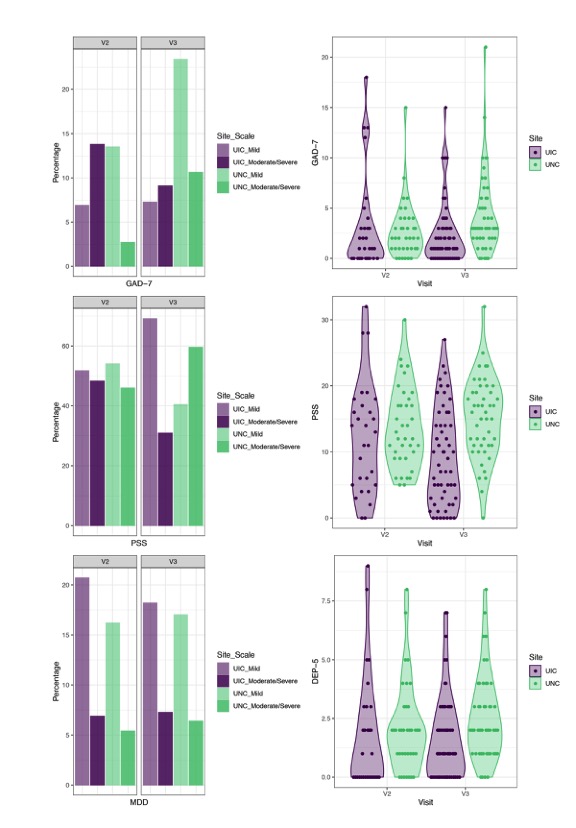

**Fig. S1. Study design and participant characteristics.** a) GAD-7; b) PSS and c) MDD percentage and d) GAD-7, e) PSS and f) DEP-5 scores by visit (v2, 24 GWs; v3, 34 GWs), by site (Chicago, red; Chapel Hill, blue), and by category (mild; moderate/severe). There are statistically significant differences at their 3^rd^ trimesters between Chicago and Chapel Hill for GAD-7 and DEP-5 scores (p=0.02) and PSS scores (p=0.0004). Similar trends were also observed in PSS score independently of the trimester (p=0.003). MDD was assessed by PHQ-9 in Chicago or EPDS in Chapel Hill.

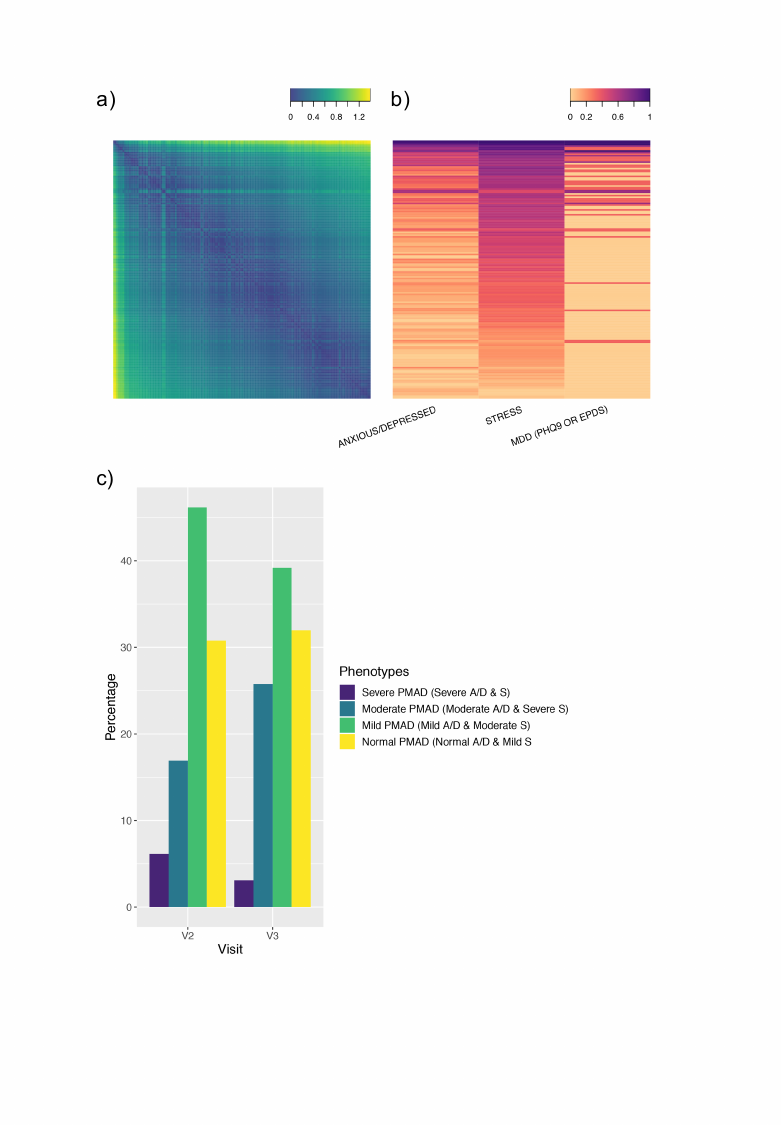

**Figure S2. Factor analysis and participant clustering based total scores of GAD-7, PSS and DEP-5.** a) correlation between the mental health questionnaire scores; b) clustering of total scores; c) associated values for each participant for mental health questionnaire; d) percentage of participants in each cluster by trimester. No statistically significant differences by trimesters (p>0.1)

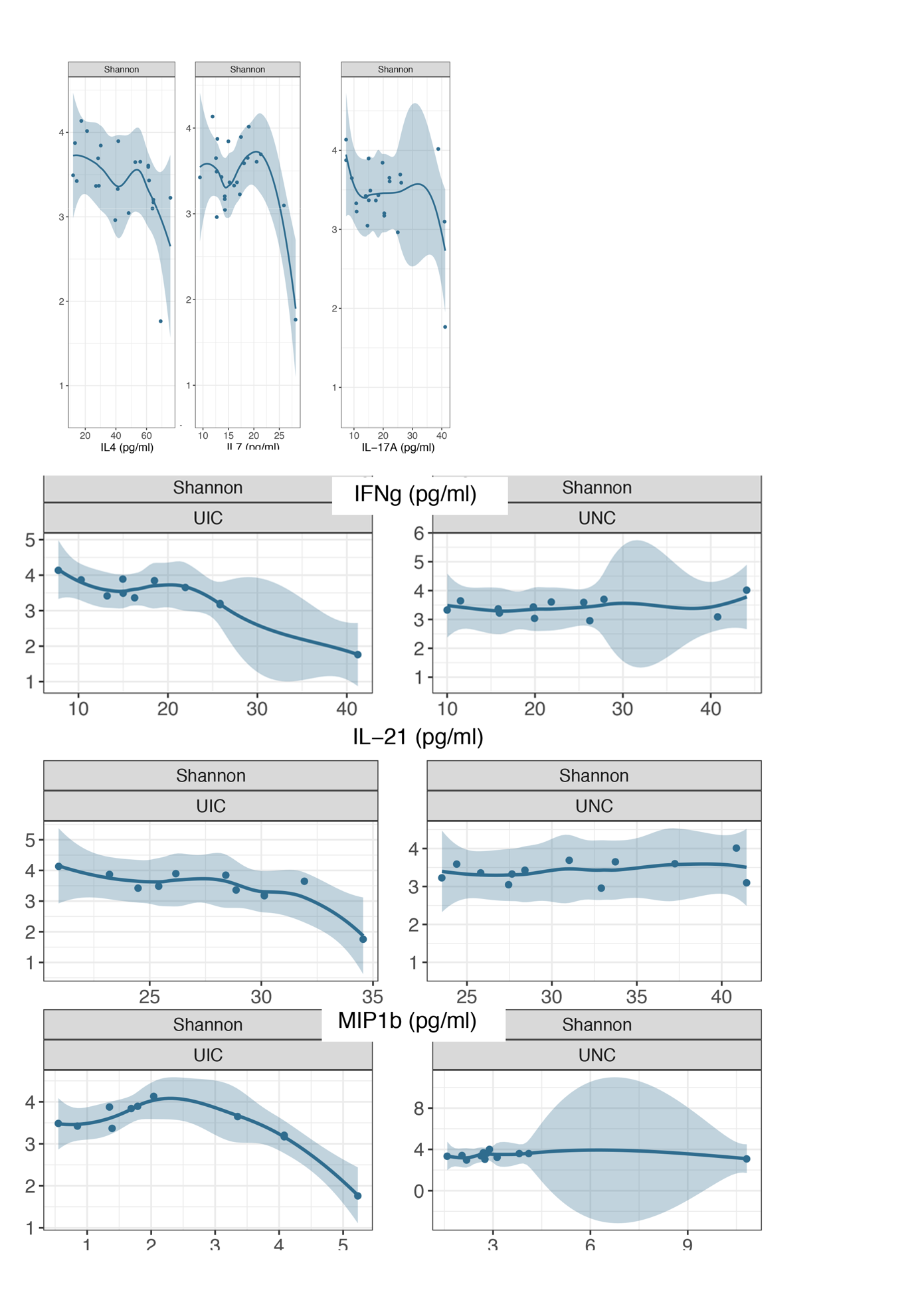

**Fig. S3. Alpha diversity measured by Shannon index was associated with the concentration of several cytokines in blood, i.e., IL-4, IL-7 and IL-17A, and some of them were site dependent (IFN-g, IL-21, MIP1b). IL-4, IL-7 and IL-17A** were associated with decreased alpha-diversity independently of the site and the participant’s gestational weeks (fdr-p<0.01), and towards signficicance when stratified by recruitment site (fdr-p<0.05) . IFN-g was associated with Shannon diversity (fdr-pvalue=0.005) just for the Chicago recruitment site. The rest of the cytokines showed a trend towards significance (fdr-pvalue<0.05). IFN-g, IL-21 and MIP1b when PERMANOVA calculations were blocked by Site, they appeared to trend towards significance (fdr-pvalue<0.05). Note that the IFN-g then became not significant at that point.

**Fig. S4. Beta-diversity estimated by Unifrac showed trends toward significance with several socio-demographic factors.** PERMANOVA calculations were adjusted by Site.

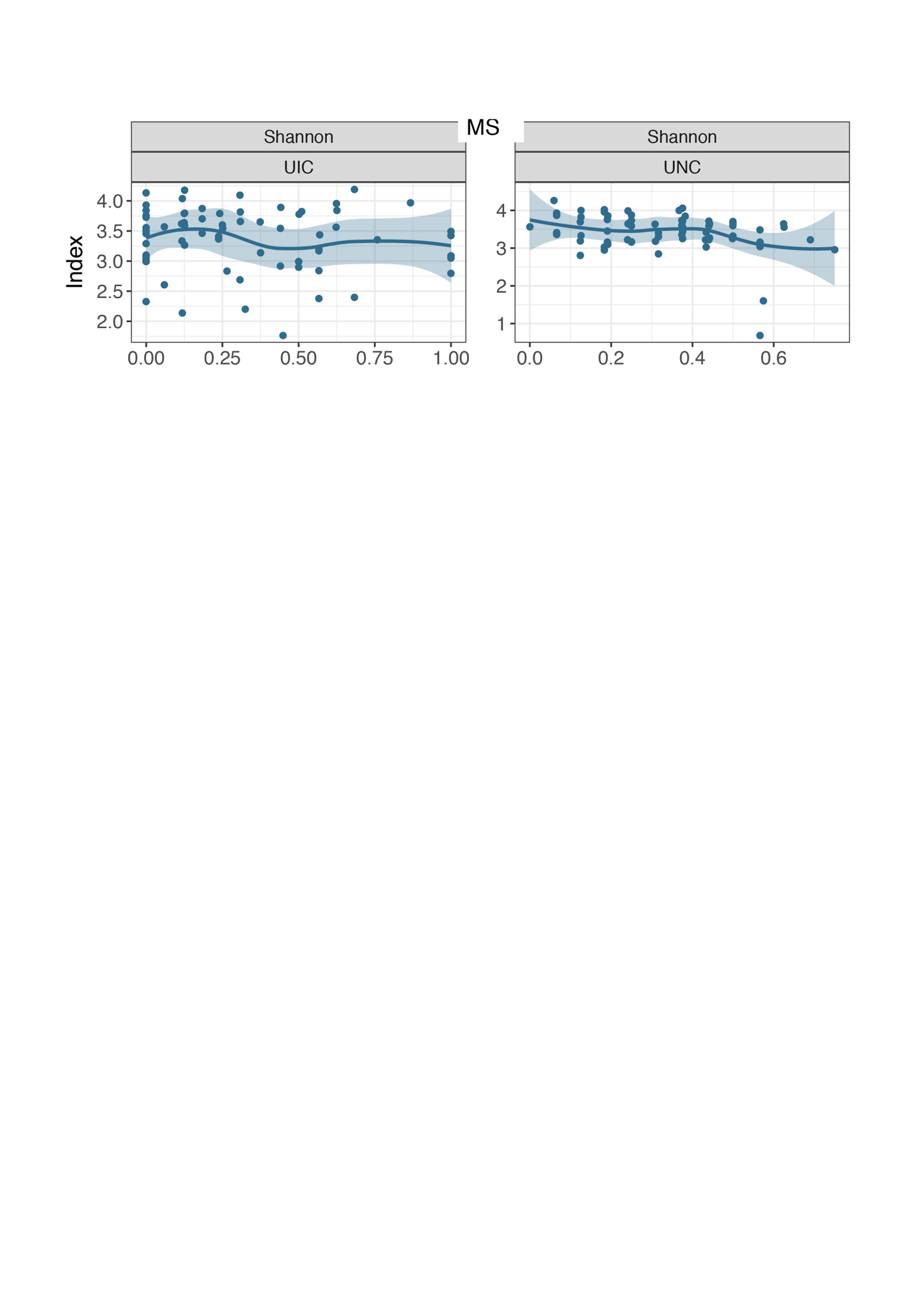

**Fig. S5. Alpha diversity measured by Shannon index was slightly associated with the mixed state factor, MS, fdr-pvalue=0.013, when stratified by recruitment site.** None of the other factors were associated with alpha-diversity.

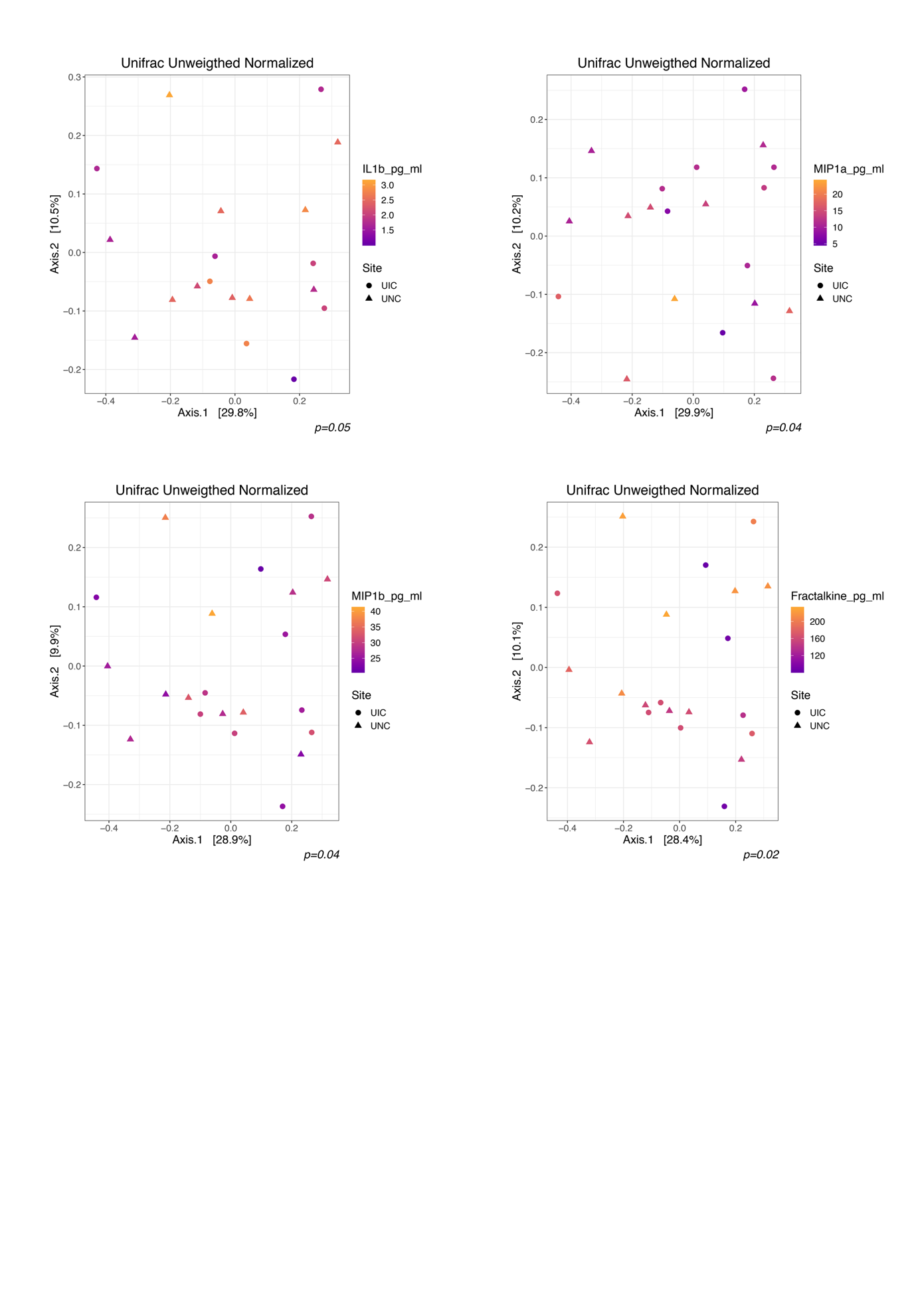

**Fig. S6. Beta-diversity estimated by Unifrac showed trends toward significance with several cytokines.** PERMANOVA calculations were adjusted by Site.

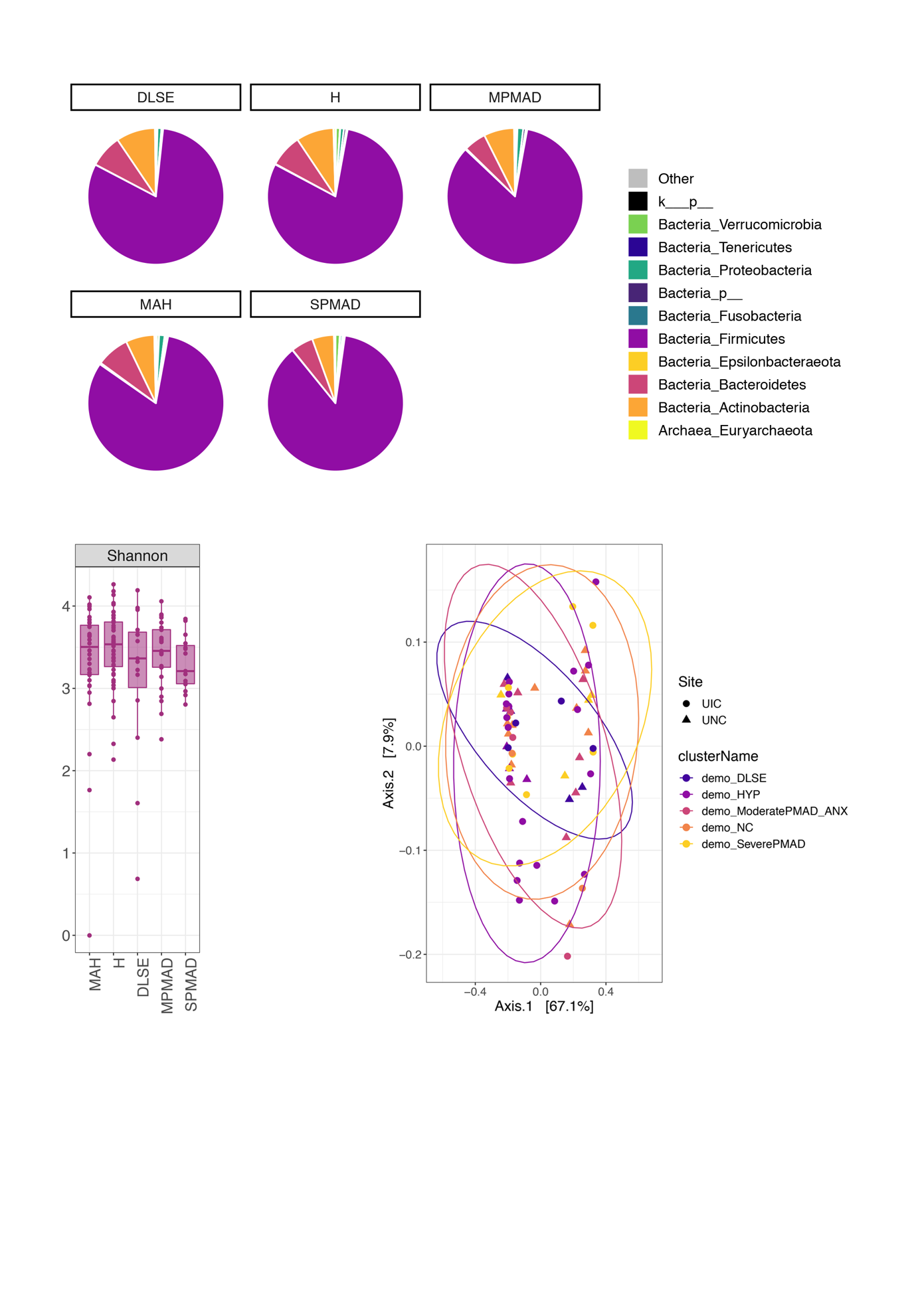

**Fig. S7. Abundance, alpha and beta diversity for each of the identified phenotypes**

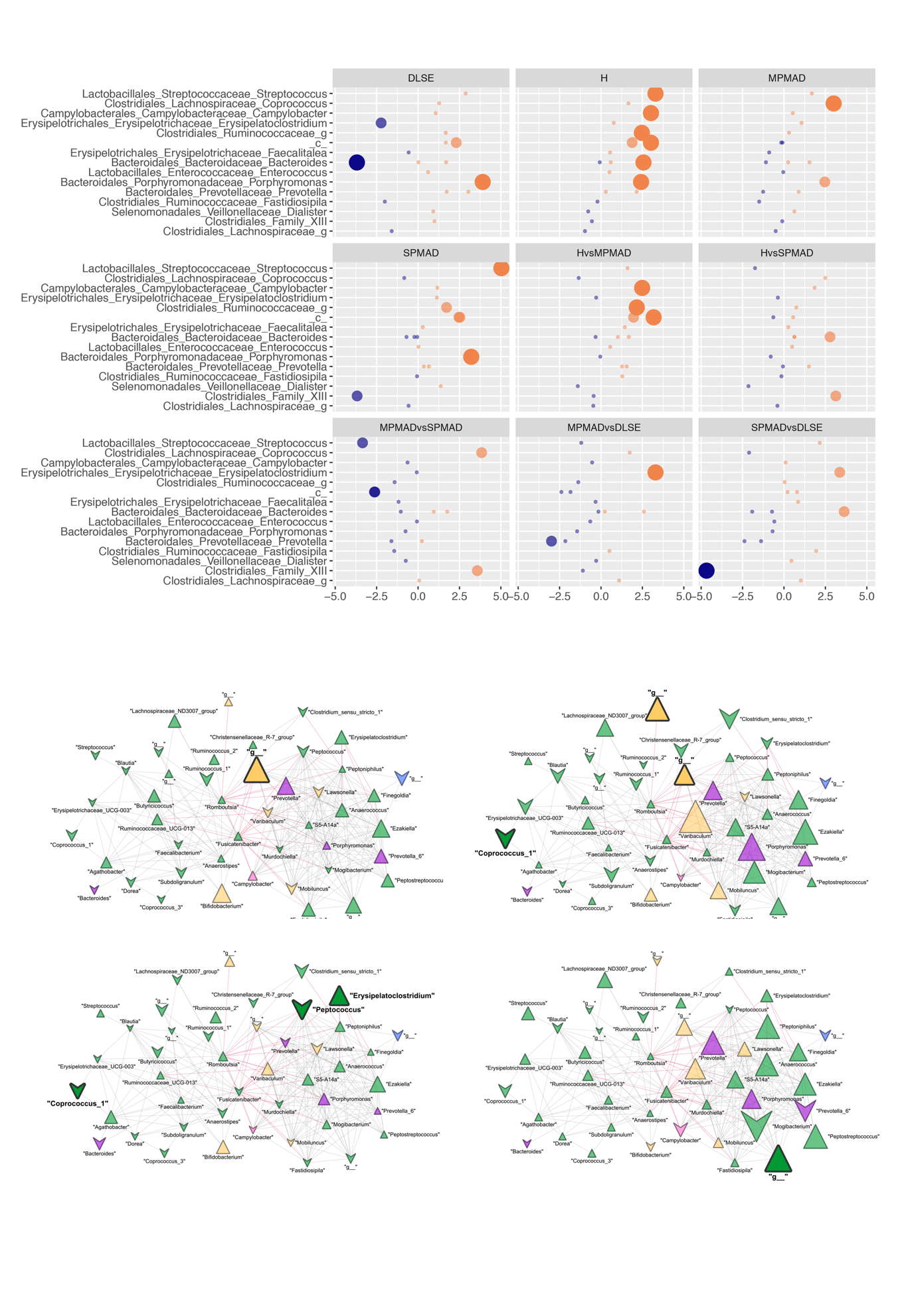

**Fig. S8. Co-abundance networks for the H, DLSE, MPMAD and SPMAD compared with the MAH cases.** Size is related to the fold change between the normalize abundance at the genus levels. Triangles indicate that the abundance was higher in the given phenotype than in the control. Bolded nodes indicate that the difference in advance was significant at a fdr-corrected p-value<0.1. Red edges indicate a negative co-abundance association between 2 given nodes. Only edges that have a correlation greater than 0.25 and a frd-corrected p-value<0.001.

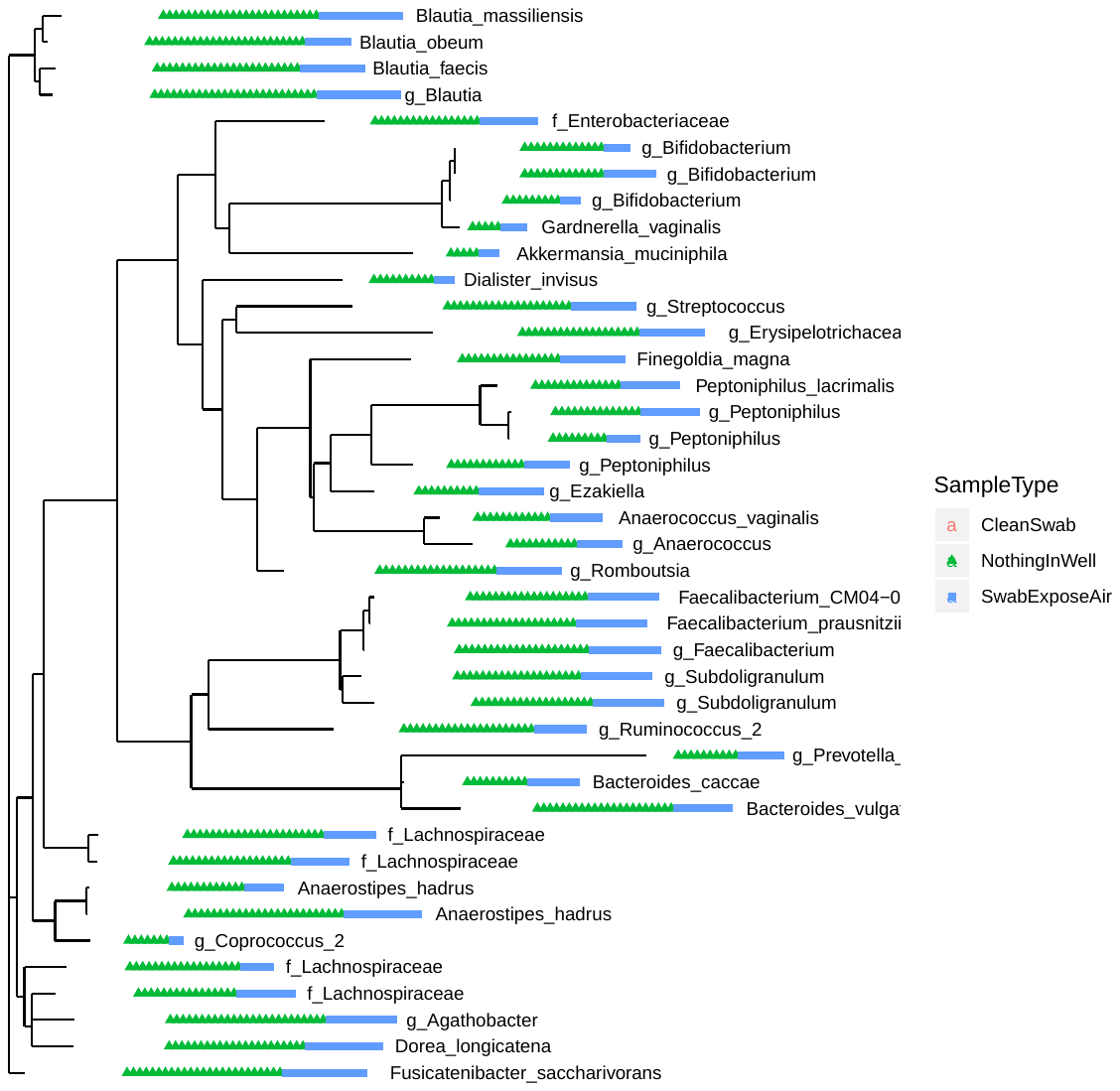

**Figure S9. Contamination in the control wells.** The ESVs summarized at the genes level that were present in the control samples are depleted here. The number of dots indicate the number of samples that each genus was present for each of the control types.

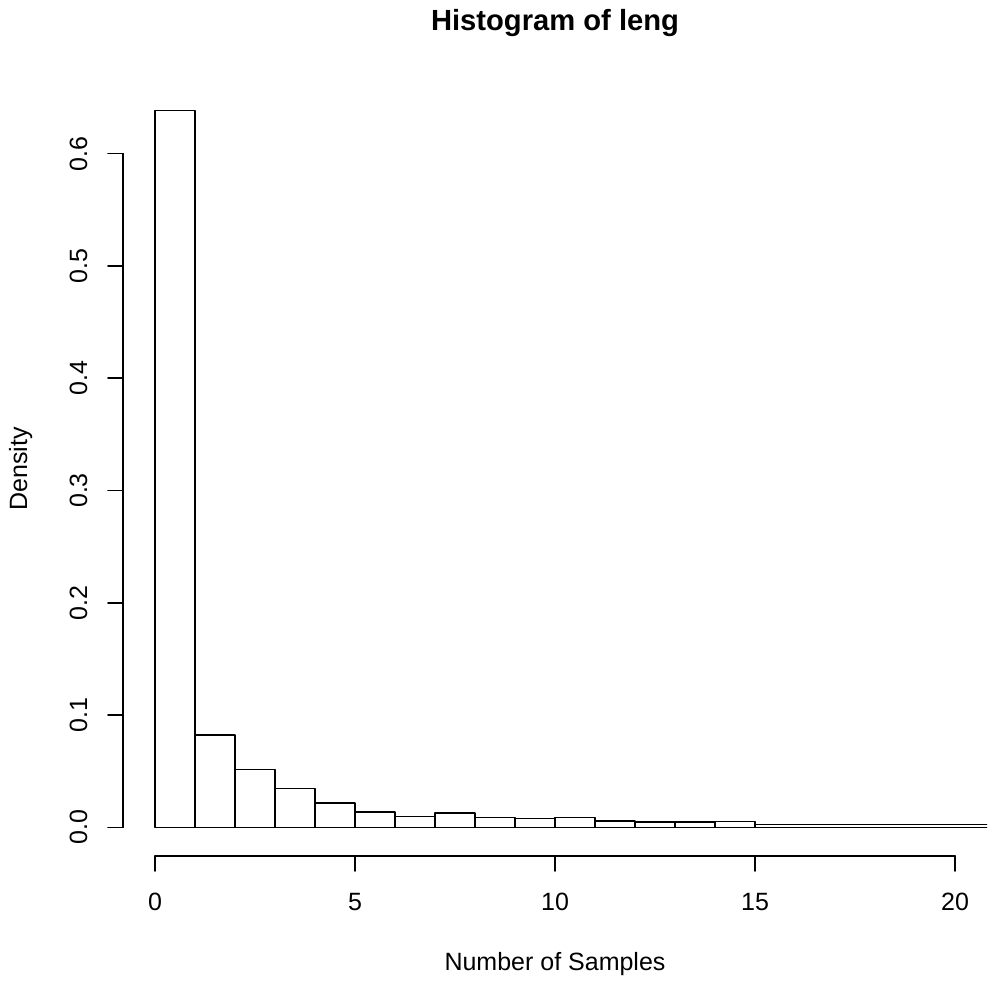

**Figure S10. Histogram of the number of times each ESV is repeated per sample.** We assumed that ESVs that were present in more than 15 samples (90% of the total samples) were no contaminants.
